# Compassionate Use of REGEN-COV^®^ in Patients with COVID-19 and Immunodeficiency-Associated Antibody Disorders

**DOI:** 10.1101/2021.11.05.21265911

**Authors:** David Stein, Ernesto Oviedo-Orta, Wendy A Kampman, Jennifer McGinniss, George Betts, Margaret McDermott, Beth Holly, Johnathan M Lancaster, Ned Braunstein, George D Yancopoulos, David M Weinreich

## Abstract

**Background:** Patients with immunodeficiency-associated antibody disorders are at a higher risk of prolonged/persistent COVID-19 infection, having no viable treatment options.

**Methods:** This is a retrospective analysis of patients with primary and/or secondary immunodeficiency-associated antibody disorders who received casirivimab and imdevimab (REGEN-COV^®^) under emergency compassionate use. The objectives were to describe safety and response to REGEN-COV, with a focus on the subset of patients who had COVID-19 duration ≥21 days prior to treatment. Quantitative (change in oxygenation status and/or viral load) and/or qualitative (physician-reported clinical status) patient outcomes data are reported.

**Results:** Outcome data are available from 64 patients who received REGEN-COV. Improvement in ≥1 outcome measure was observed in 90.6% of the overall patient group. Thirty-seven of these patients had COVID-19 duration ≥21 days prior to treatment, with a median time from RT-PCR diagnosis to REGEN-COV administration of 60.5 days. Of the 29 patients with COVID-19 duration ≥21 days prior to treatment who had available outcome data, 96.6% showed improvement in ≥1 outcome measure evaluated following use of REGEN-COV. In the 14 patients who had post-treatment RT-PCR results available, 11 (78.6%) reported a negative RT-PCR following treatment with REGEN-COV, with 5 patients (45.5%) reporting a negative RT-PCR within 5 days of treatment and 8 (72.7%) reporting a negative RT-PCR within 21 days of treatment.

**Conclusions:** In this retrospective analysis of immunodeficient patients who were granted REGEN-COV under the compassionate use program, REGEN-COV treatment was associated with rapid viral clearance and clinical improvement in the evaluable patients with long-standing COVID-19.

**Summary:** Patients with immunodeficiency-associated antibody disorders are at a higher risk of prolonged/persistent COVID-19 infection. In this retrospective analysis, compassionate use of REGEN-COV in such patients was associated with rapid viral clearance and/or clinical improvement in the evaluable patients.

## INTRODUCTION

Severe acute respiratory syndrome coronavirus 2 (SARS-CoV-2), the causal agent of coronavirus disease 2019 (COVID-19), first emerged in 2019, and has led to a global pandemic. Patients infected with SARS-CoV-2 are at risk of developing a range of respiratory conditions, from mild to severe symptoms, and sometimes fatal illness [1]. Patients with either primary or secondary immunodeficiency-associated antibody disorders that have an inherited or acquired inability to produce antibodies against infective agents have a high risk of suffering from prolonged or persistent SARS-CoV-2 infection, increasing their morbidity and mortality risk [2–4]. These patients are also likely to show lack of, or significantly less effective, protective immune responses to vaccines [5], and may have to rely on alternate prevention and treatment regimes.

The treatment of immunocompromised patients with COVID-19 using convalescent plasma is controversial due to lack of specificity and variable content of potent neutralizing antibodies [6–8]. In February 2021, the United States Food and Drug Administration issued a revision of the Emergency Use Authorization for COVID-19 convalescent plasma, for the treatment of hospitalized patients with COVID-19 who have impaired humoral immunity and cannot produce an adequate antibody response [9]. However, results from 3 small randomized controlled trials in patients with moderate-to-severe COVID-19 showed no significant differences in improved clinical outcomes between the convalescent plasma-treated and placebo groups [10–12].

Casirivimab and imdevimab (REGEN-COV^®^) is a combination of 2 human, immunoglobulin G1 monoclonal antibodies (mAbs) that bind the receptor-binding domain of the SARS-CoV-2 spike protein and block interaction with angiotensin-converting enzyme 2 [13]. REGEN-COV targets separate non-overlapping epitopes thereby neutralizing SARS-CoV-2, with a reduced likelihood of viral escape due to genetic mutations [14]. Furthermore, this combination of mAbs has been shown to retain neutralization potency against multiple SARS-CoV-2 variants [14]. The blood levels of SARS-CoV-2 neutralizing activity that are achieved with REGEN-COV are approximately 10,000-to 100,000-fold higher than that achievable with high-titer convalescent plasma. Correspondingly, REGEN-COV has demonstrated convincing activity across multiple phase 2 and 3 studies and across the continuum of the disease, including pre- and post-exposure prophylaxis [15], as well as in the treatment of both outpatients and hospitalized patients [16–19]. Across these studies, the greatest benefit was observed in patients who had not yet developed their own innate immune response at baseline (ie, who did not make their own anti-SARS-CoV-2 antibodies and are thus termed “seronegative”); such high-risk seronegative patients may share characteristics with individuals with an immunodeficiency who cannot mount an antibody response.

Informed by the clinical data from REGEN-COV program, Regeneron has, with the approval of the FDA, been granting compassionate use to patients with COVID-19 and primary and/or secondary immune deficiencies. This retrospective analysis of such patients who received REGEN-COV under emergency compassionate use gathered data to provide a better understanding of clinical outcomes in this vulnerable group. While data for all patients (the overall patient group) are presented, we give particular focus to the subset of patients with COVID-19 duration ≥21 days prior to treatment, as this subset reflects those unable to effectively clear the virus.

## METHODS

### Analysis Overview

This is a retrospective, descriptive data analysis using de-identified patient data from patients who received REGEN-COV under emergency compassionate use from 2 September 2020 to 29 March 2021.

Patients were evaluated and eligible for REGEN-COV under compassionate use if they had a serious, life-threatening disease (where the patient cannot wait for the usual approval process), had sufficient evidence of a positive risk/benefit of using the experimental agent for the condition affecting the patient, were not eligible for any available clinical trial, and had no viable or available treatment options. To be eligible for compassionate use, patients had to have a positive SARS-CoV-2 reverse transcription polymerase chain reaction (RT-PCR) within approximately 1 week of treatment. Each request for compassionate use was reviewed by the compassionate use committee consisting of senior leaders at Regeneron Pharmaceuticals, Inc. prior to submission to the FDA for approval. Requesting physicians in the US had to obtain authorization from FDA for emergency IND and comply with the requirements of 21 CFR 312.310; ex-US requests had to comply with all local regulations. Further information and details about the compassionate use program can be found at https://www.regeneron.com/downloads/regeneron-compassionate-use-request.pdf.

### Outcome Measures

The objective of this retrospective analysis was to describe safety and response to REGEN-COV in patients with primary and/or secondary immunodeficiency-associated antibody disorders that were evaluated and approved for drug use under compassionate use. Response to REGEN-COV is described using quantitative patient outcome data (change in oxygenation status and/or viral load) and/or qualitative patient outcome data (physician-reported clinical status). Serious adverse events and death were also evaluated.

Oxygenation status is described as oxygen requirement and correlated oxygen saturations. Viral load is described by either SARS-CoV-2 RT-PCR status (negative or positive) and/or cycle threshold (Ct) values in RT-PCR, prior to and after compassionate use of REGEN-COV. Descriptive safety data are also presented.

### Ethics

The treating physician was responsible for compliance in accordance with the principles of the Declaration of Helsinki, International Council for Harmonisation Good Clinical Practice guidelines, and applicable regulatory requirements for administering an investigational product under compassionate use. The treating physician was also responsible for all safety reporting and regulatory obligations associated with the conduct of the compassionate use as required by applicable law. The institutional review board (WCG IRB, Puyallup, WA; IRB tracking number 20212896) found that this research met the requirements for a waiver of consent under 45 CFR 46 116(f) and 45 CFR 46.116(d). All patients provided written informed consent before receiving treatment.

## RESULTS

### Patients with COVID-19 Duration ≥21 Days Prior to Treatment

In total, 174 patients from the US were evaluated and approved to receive REGEN-COV under the emergency compassionate use program as of 29 March 2021, of whom 95 (54.6%) had primary and/or secondary immunodeficiency-associated antibody disorders. Of these patients, 85 received intravenous REGEN-COV and data is available for 64 of these patients. Of the 64 patients with available data, 37 (43.5%) had COVID-19 duration ≥21 days prior to treatment.

In the subset of patients with COVID-19 duration ≥21 days prior to treatment, 64.9% were male, the mean age was 49.1 years (median: 52.0 years; min–max: 1– 75), 27.0% were aged ≥65 years, and 27/37 (73.0%) were inpatients. The median time from RT-PCR diagnosis to administration of REGEN-COV was 60.5 days (min– max: 25–218; mean: 83.2 days). Three patients (8.1%) had primary immunodeficiency-associated antibody disorders, 34 patients (91.9%) had secondary causes of immunodeficiency-associated antibody disorders (malignant or drug induced); the most common causes included treatment with anti-CD20 (rituximab), acute lymphocytic leukemia, follicular lymphoma, diffuse B-cell lymphoma, and other non-Hodgkin’s lymphomas (**Table 1**).

**Table 1.**
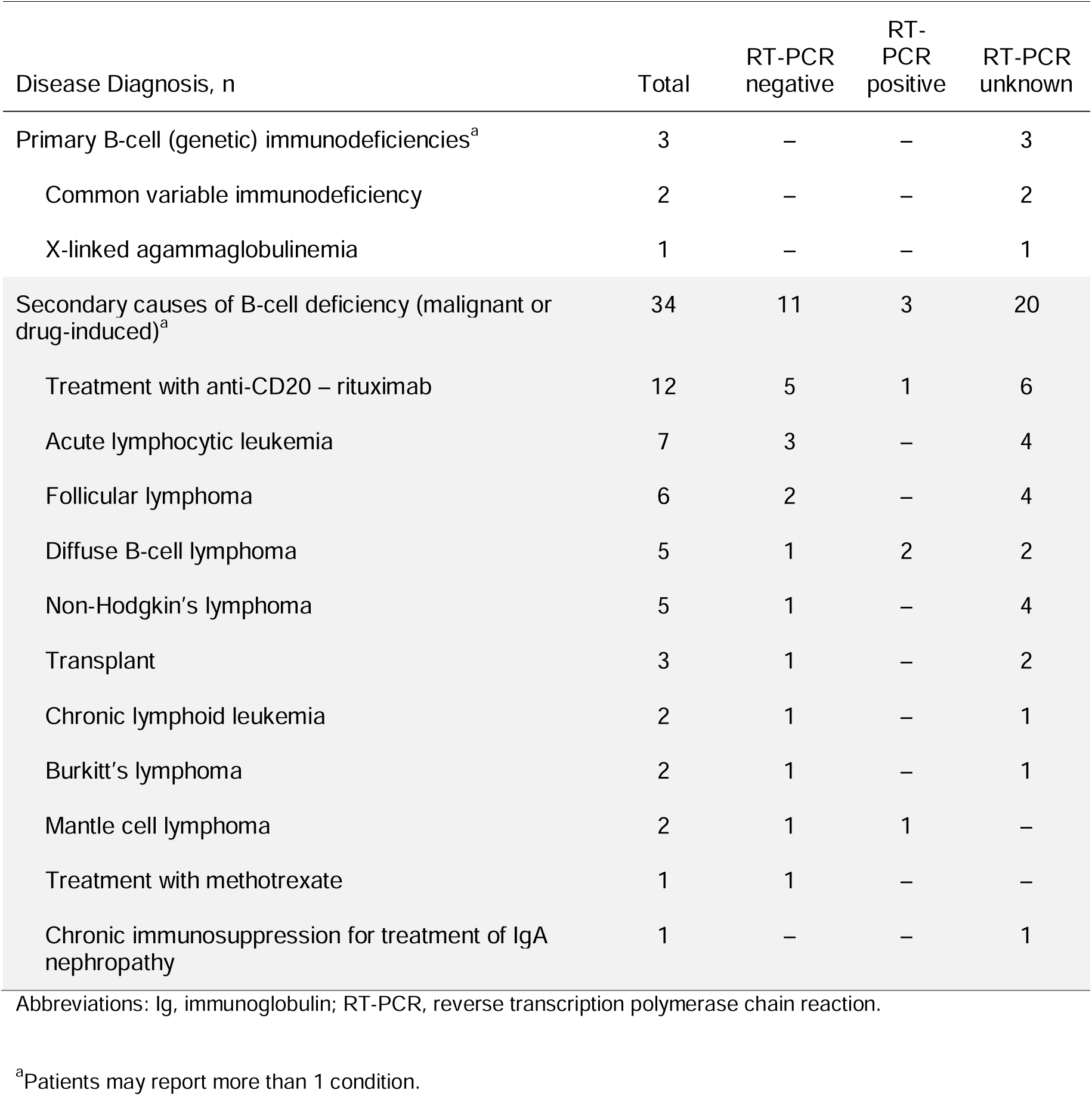
RT-PCR outcomes by disease diagnosis in the subset of patients with disease duration ≥21 days prior to treatment

Of the 37 patients with COVID-19 duration ≥21 days prior to treatment, qualitative or quantitative outcome data were available for 29 patients: 23 (79.3%) had qualitative physician assessment at follow-up, 22 (75.9%) had oxygenation status measured, and 14 (37.8%) had an evaluable RT-PCR measurement following REGEN-COV treatment. Overall, 96.6% of patients (28/29; 95% confidence interval [CI]: 80.4–99.8) showed improvement in 1 or more of these measures following compassionate use of REGEN-COV. Of the 23 patients who had a qualitative physician assessment at follow-up, 22 (95.7%; 95% CI: 76.0–99.8) had improved. Of the 22 patients who had oxygenation status measured at follow-up, all 22 (100%; 95% CI: 81.5–100) had an improved oxygenation status, either increased oxygen saturation or reduced oxygen requirement (**Figure 1**). Of the 14 patients who had an evaluable RT-PCR measurement during follow-up, 11 (78.6%) reported a negative RT-PCR following treatment with REGEN-COV (**Figure 2**); the 3 patients who remained RT-PCR positive were tested on average of 4 days (min–max: 1–9) post-treatment with REGEN-COV. Of the 11 patients who became RT-PCR-negative, 5 patients (45.5%) developed a negative RT-PCR within 5 days of treatment and 8 (72.7%) developed a negative RT-PCR within 21 days of treatment (**Table 1**). Of note, these 14 patients had a mean time since RT-PCR diagnosis prior to treatment of 62.8 days (median 53.5; range 25–215; **Figure 2**).

**Figure 1.**
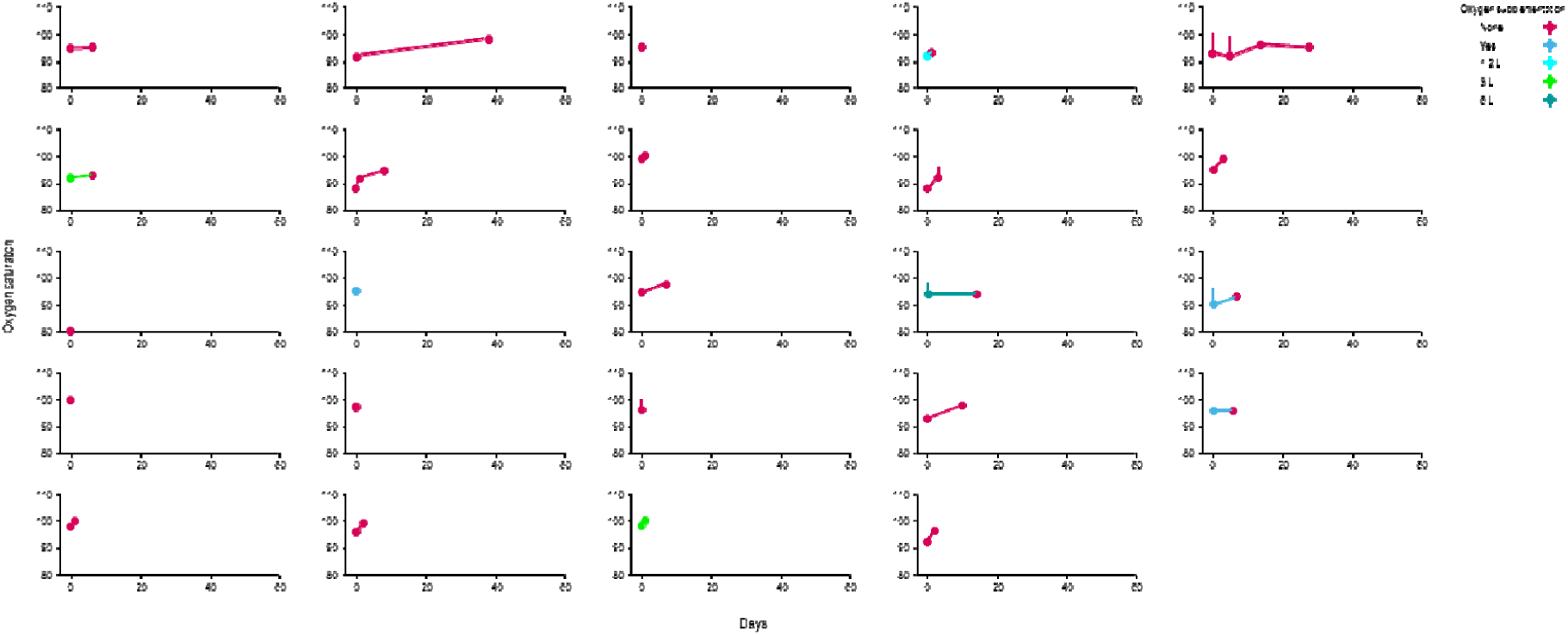
Change in oxygenation status over time in patients with primary and/or secondary immunodeficiencies in the subset of patients with COVID-19 duration ≥21 days prior to treatment Treatment was administered on day 0. Oxygen saturation over time (days) in (A) patients with no supplemental oxygen at baseline, and (B) those patients with some supplemental oxygen at baseline. The volume of oxygen supplementation is indicated by color, and shows increases and decreases in supplementation.

**Figure 2.**
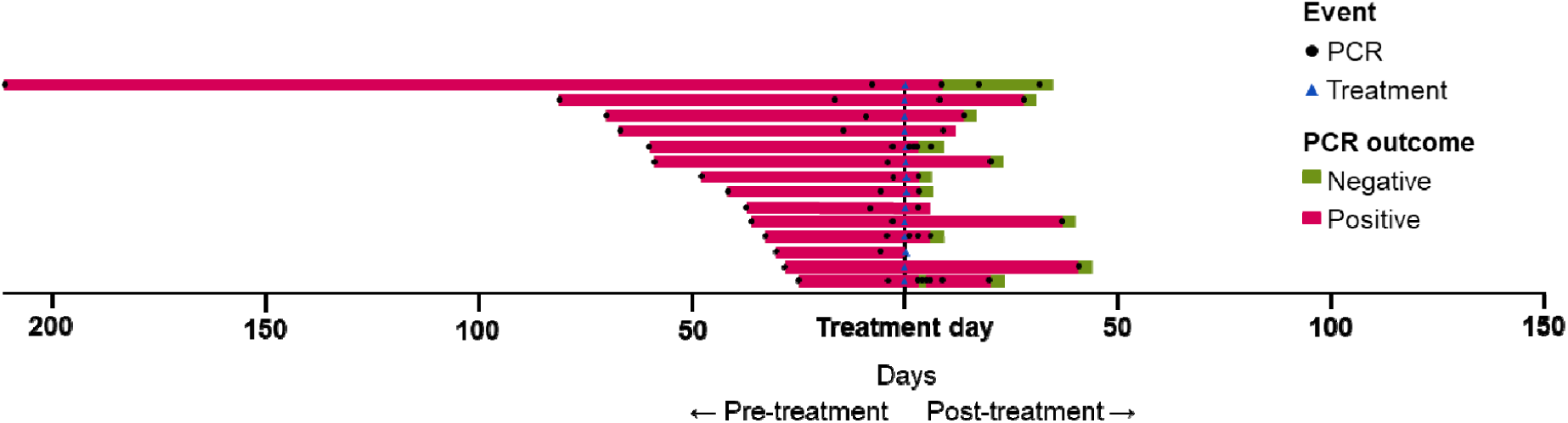
Time from RT-PCR diagnosis to compassionate use of REGEN-COV to post-baseline RT-PCR where available in patients with COVID-19 duration ≥ 21 days prior to treatment Abbreviations: COVID 19, coronavirus disease 2019; RT-PCR, reverse transcription polymerase chain reaction. Circles indicate days on which patients received a RT-PCR test. The time between first RT-PCR test leading to a diagnosis of COVID-19 and treatment varied significantly, with some receiving treatment within a couple of days but others waiting over 200 days. Following traetment (indicated with blue triangle), patients underwent subsequent RT-PCR tests: green bars indicate a negative test result and pink indicates a positive test result.

### Overall Patient Group

In the overall patient group, 62.4% were male, the mean age was 47.4 years (median: 52.0 years; min–max: 1–79), 24.7% of patients were aged ≥65 years, and the majority were inpatients (65/85; 76.5%). The median time from RT-PCR diagnosis to administration of REGEN-COV was 13.0 days (min–max: 1–218; mean: 40.8 days). Thirteen patients (15.3%) had primary (genetic) immunodeficiency-associated antibody disorders, and 72 (84.7%) had secondary causes of immunodeficiency-associated antibody disorders (malignant or drug-induced) (**Supplementary Table 1**).

Qualitative or quantitative outcome data were available for 64/85 patients: 58/64 (90.6%) had qualitative physician assessment at follow-up; 39/64 (60.9%) had oxygenation status measured; and 14/64 (21.9%) had Ct measured, approximately 1 week prior to and after compassionate use of REGEN-COV. Overall, 90.6% of patients (58/64; 95% CI: 80.1–96.1) showed improvement in 1 or more of these measures following compassionate use of REGEN-COV. Qualitative physician follow-up showed that 52/58 patients (89.7%; 95% CI: 78.2–95.7) had improved. A total of 33/39 patients (84.6%; 95% CI: 68.8–93.6) had an improved oxygenation status, either increased oxygen saturation or reduced oxygen requirement (**Supplementary Figure 1**) and 14/14 of patients (100%; 95% CI: 89.1–100) had improved Ct on a post-baseline RT-PCR (**Figure 3**).

All 14 patients with a quantitative RT-PCR assessment showed rapid reduction in viral load with REGEN-COV treatment despite having a diagnosis of immunodeficiency (as shown by an increase in cycle time in **Figure 3**). Qualitative RT-PCR was evaluable in 28 patients from the overall patient group, and 20 (71.4%) of these reported a negative RT-PCR post-baseline; 3 additional patients had a fluctuating negative RT-PCR test. RT-PCR outcomes by disease diagnosis in the overall patient group are summarized in **Supplementary Table 1**.

**Figure 3.**
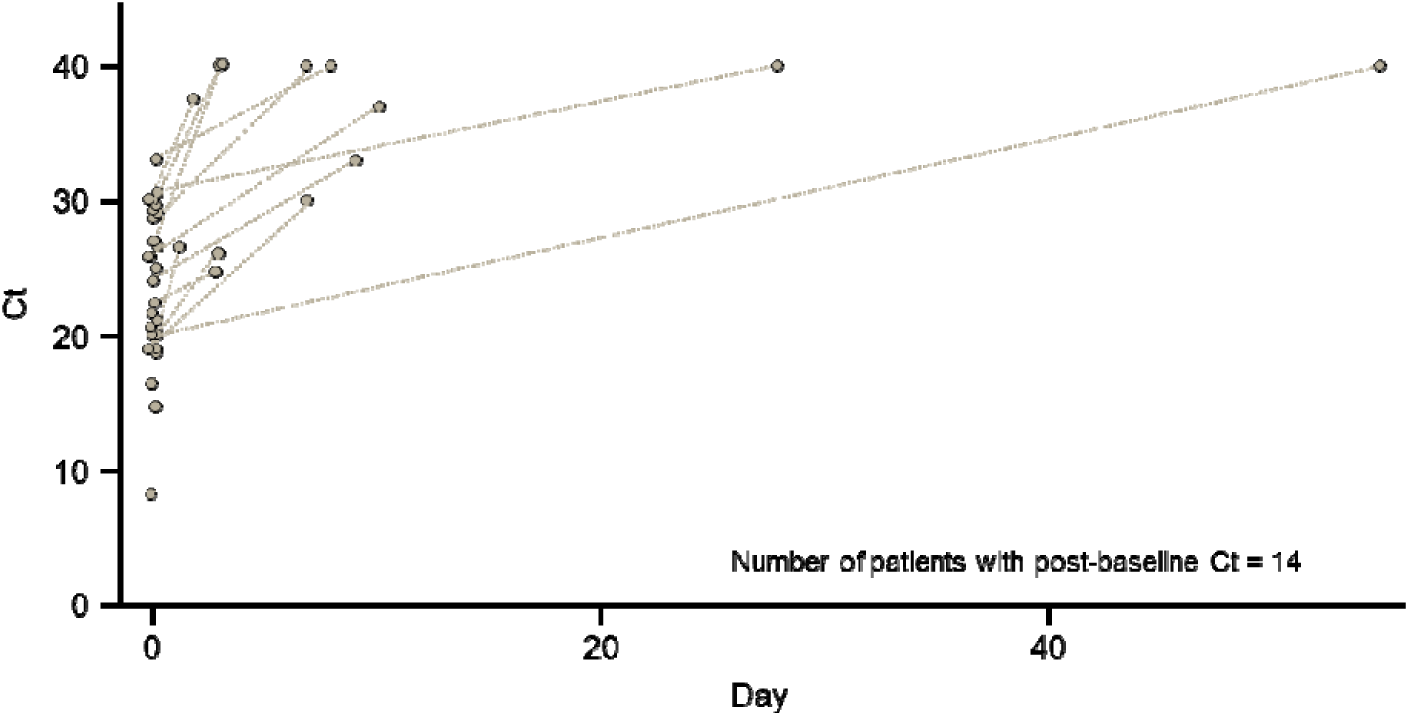
Change in Ct over time in patients with primary and/or secondary immunodeficiencies in the overall patient group Abbreviations: Ct, cycle threshold; RT-PCR, reverse transcription polymerase chain reaction. Treatment was administered on day 0. Ct values are from the most recent RT-PCR prior to treatment. Viral load was determined by Ct values from RT-PCR. Ct values in 14 patients post-baseline showed an improvement correlating with a reduction in viral load after receiving REGEN-COV. Note: Ct not detected was assumed to have a value of 40.

### Safety

In the overall patient group, 10/85 patients (11.8%) experienced serious adverse events (SAEs; as defined by physician-completion of an SAE form), of which only 1 event was an infusion-related reaction judged as possibly related to REGEN-COV. Two patients who experienced SAEs had a COVID-19 duration ≥21 days prior to treatment. There were 2 deaths reported, neither of which were attributed by the investigator to treatment with REGEN-COV: 1 from hypoxia; and 1 from respiratory failure secondary to chronic COVID-19 and diffuse alveolar hemorrhage complicated by pulmonary aspergillosis. The events reported were consistent with COVID-19 and its associated complications and due to patients’ concurrent medical conditions.

## DISCUSSION

Because immunocompromised patients cannot generate their own antibodies against SARS-CoV-2, whether by vaccination or natural infection, they represent a particularly vulnerable and high-risk population. Antibody-related treatments for SARS-CoV-2 – compensating for the lack of endogenous antibodies – are potentially viable treatment options, including polyclonal antibodies such as convalescent plasma [20]. However, the available clinical data have shown no efficacy in patients with COVID-19 when treated with convalescent plasma [10–12], which generally has limited amounts of SARS-CoV-2 neutralizing antibody levels. In contrast, 10,000– 100,000-fold higher levels of neutralizing activity can be achieved with recombinant antibody treatments such as REGEN-COV, which has demonstrated convincing activity across multiple phase 2 and 3 studies and across the continuum of the disease, including in high-risk seronegative patients who share some characteristics with individuals with an immunodeficiency who cannot mount an antibody response [15–19].

Case reports have demonstrated treatment benefit with REGEN-COV in individual patients with primary and/or secondary immunodeficiencies, either alone or in combination with standard-of-care antiviral therapies [21–25]. Furthermore, in a report of 25 solid organ transfer outpatients with mild-to-moderate COVID-19, no patient experienced symptom progression or required hospitalization following treatment with REGEN-COV [24]. Additionally, in a case report of a kidney transplant recipient with COVID-19 following poor response to vaccination, treatment with REGEN-COV led to seroconversion and improved clinical and virologic outcomes [26].

As far as we are aware, this is the first report describing outcomes following REGEN-COV treatment for COVID-19 in a series of patients with primary and/or secondary immunodeficiency-associated antibody disorders. In this retrospective analysis, 96.6% of patients with COVID-19 duration ≥21 days prior to treatment showed marked improvement in 1 or more qualitative and/or quantitative outcomes following compassionate use with REGEN-COV, with 85.7% showing improvement within 7 days of treatment. Most patients with evaluable RT-PCR data during follow-up showed rapid reductions in viral load, despite many having prolonged duration of infection prior to administration of REGEN-COV.

This case series provides data which are consistent with the totality of the data from phase 2 and 3 studies of REGEN-COV [15–19], in which the greatest benefit with REGEN-COV was seen in high-risk patients who had not yet mounted their own antibody response, and thus may share some characteristics consistent with immunodeficient patients. In those studies, REGEN-COV treatment in those who had not yet mounted their own immune response was associated with robust and rapid reduction in viral load as well as improvement in clinical outcome measures. The data from this case series describe high recovery rates comparable to those in the phase 2 and 3 studies who had not yet mounted an immune response.

In some territories, REGEN-COV is approved for the treatment and prevention of COVID-19 broadly. However, in the United States the current authorization for treatment with REGEN-COV only covers recently diagnosed patients (within 10 days of symptom onset) [27] and thus would not allow for treatment of these immunocompromised patients with long-standing disease or who are hospitalized. In this retrospective analysis of COVID-19 patients with primary and/or secondary immunodeficiency-associated antibody disorders who were granted REGEN-COV under the compassionate use program, the majority showed rapid clinical improvement and viral clearance following treatment with REGEN-COV. These data, along with the previously reported clinical trial data, support the broader use of REGEN-COV for the treatment and prevention of COVID-19 in the immunocompromised patient population.

## Supporting information

Author Lancaster COI

Author McDermott COI

Author Braunstein COI

Author Kampman COI

Author Holly COI

Author Weinreich COI

Author Stein COI

Author Orta COI

Author Betts COI

Author Yancopoulos COI

Author McGinniss COI

## Data Availability

Qualified researchers may request access to study documents (including the clinical study report, study protocol with any amendments, blank case report form, and statistical analysis plan) that support the methods and findings reported in this manuscript. Individual anonymized participant data will be considered for sharing once the indication has been approved by a regulatory body, if there is legal authority to share the data and there is not a reasonable likelihood of participant re-identification. Submit requests to https://vivli.org/.

## Notes

### Funding

This work was supported by Regeneron Pharmaceuticals, Inc.

### Conflict of Interest

DS, EO-O, WAK, JM, GB, MM, BH, JML, NB, and DMW are Regeneron employees/stockholders. GDY is a Regeneron employee/stockholder and has issued patents (U.S. Patent Nos. 10,787,501, 10,954,289, and 10,975,139) and pending patents, which have been licensed and receiving royalties, with Regeneron.

## Acknowledgements

The authors would like to thank the patients, their families, and the physicians involved in this work, as well as Caryn Trbovic, PhD, from Regeneron Pharmaceuticals for medical writing support; and Prime, Knutsford, UK, for formatting and copy-editing suggestions and for assistance with development of the manuscript.

## Notes

### Competing Interest Statement

ICMJE disclosure forms provided by the authors are available with the full text of this article.

### Author Declarations

The institutional review board (WCG IRB, Puyallup, WA; IRB tracking number 20212896) found that this research met the requirements for a waiver of consent under 45 CFR 46 116(f) and 45 CFR 46.116(d).

