## Supplemental Materials for "Compassionate Use of REGEN-COV^®^ in Patients with COVID-19 and Immunodeficiency-Associated Antibody Disorders"

### Supplementary Appendix

Supplementary Table 1. RT-PCR outcomes by disease diagnosis in the overall patient group

| Disease diagnosis, n | Total | RT-PCR negative | RT-PCR positive | RT-PCR unknown |
| --- | --- | --- | --- | --- |
| Pure B-cell (genetic) immunodeficiencies^a^ | 13 | 2 | – | 11 |
| Common variable immunodeficiency | 5 | – | – | 5 |
| X-linked agammaglobulinemia | 3 | – | – | 3 |
| IPEX syndrome | 2 | – | – | 2 |
| IgG subclass deficiency | 2 | 1 | – | 1 |
| CTLA-4 haploinsufficiency | 1 | 1 | – | – |
| Selective IgA deficiency | 1 | – | – | 1 |
| TRNT1 genetic defect | 1 | – | – | 1 |
| Secondary causes of B-cell deficiency (malignant or drug-induced)^a^ | 72 | 17 | 8 | 47 |
| Transplant | 21 | 6 | 3 | 12 |
| Treatment with anti-CD20 – rituximab | 10 | 5 | 2 | 3 |
| Diffuse B-cell lymphoma | 8 | 1 | 2 | 5 |
| Acute myeloid leukemia | 8 | 3 | 1 | 4 |
| Acute lymphocytic leukemia | 6 | 3 | 1 | 2 |
| Non-Hodgkin’s lymphoma | 6 | 2 | – | 4 |
| T-cell lymphoblastic lymphoma/leukemia | 5 | 1 | 1 | 3 |
| Follicular lymphoma | 4 | 2 | – | 2 |
| Chronic lymphoid leukemia | 4 | 1 | – | 3 |
| Burkitt lymphoma | 3 | 1 | – | 2 |
| Mantle cell lymphoma | 2 | 1 | 1 | – |
| Lung cancer | 1 | – | – | 1 |
| Hodgkin’s lymphoma | 1 | – | – | 1 |
| Treatment with methotrexate | 1 | – | – | 1 |
| Metastatic kidney cancer | 1 | – | – | 1 |
| Synchronous primary lung cancers | 1 | – | – | 1 |
| Ovarian cancer | 1 | – | – | 1 |
| Treatment with mycophenolate mofetil (CellCept^®^, Myfortic^TM^) | 1 | – | – | 1 |
| Renal cell carcinoma | 1 | – | – | 1 |
| Malignant thymoma | 1 | – | – | 1 |
| Chronic immunosuppression for treatment of IgA nephropathy | 1 | – | – | 1 |
| Osteosarcoma metastatic to lung | 1 | – | – | 1 |

Abbreviations: CTLA-4, cytotoxic T-lymphocyte-associated protein 4; Ig, immunoglobulin; IPEX ,immunodysregulation polyendocrinopathy enteropathy X-linked; RT-PCR, reverse transcription polymerase chain reaction; TRNT1, transfer ribonucleic acid-nucleotidyltransferase 1.

^a^Patients may report more than 1 condition.

Supplementary Figure 1. Change in oxygenation status over time in patients with primary and/or secondary immunodeficiencies in the overall patient group

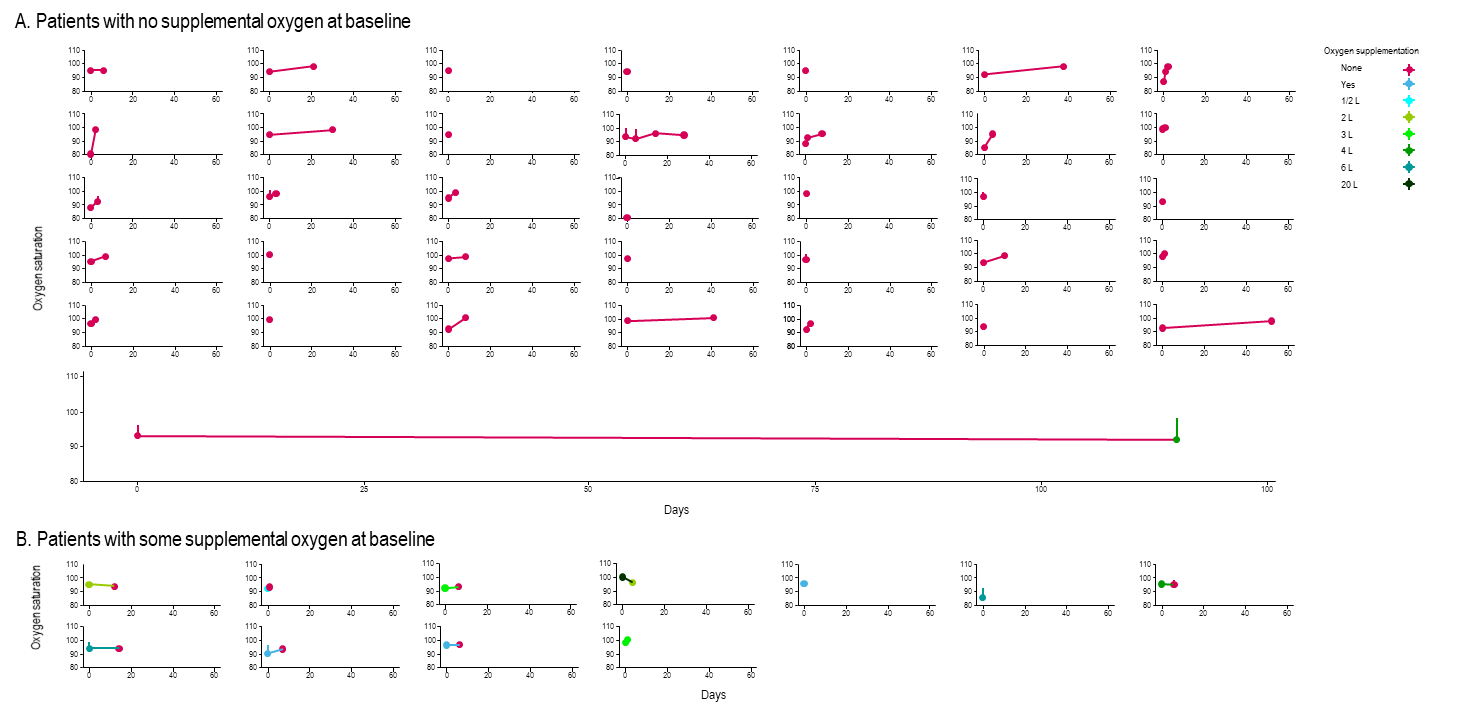

Treatment was administered on day 0. Oxygen saturation over time (days) in (A) patients with no supplemental oxygen at baseline, and (B) those patients with some supplemental oxygen at baseline. The volume of oxygen supplementation is indicated by color, and shows increases and decreases in supplementation.
